# State-of-the-Art Risk Models for Diabetes, Hypertension, Visual Diminution, and COVID-19 Severity in Mexico

**DOI:** 10.1101/2021.01.18.21250034

**Authors:** Heladio Amaya, Jennifer Enciso, Daniela Meizner, Alex Pentland, Alejandro Noriega

**Affiliations:** Prosperia, Mexico; Academy for Systems and Computation, ITD, Mexico; Biochemical Sciences, UNAM, Mexico; Retina Department, Asociación para Evitar la Ceguera en México, Mexico; MIT Media Laboratory, Massachusetts Institute of Technology, USA

## Abstract

**BACKGROUND:** Diabetes and hypertension are among top public health priorities, particularly in low and middle-income countries where their health and socioeconomic impact is exacerbated by the quality and accessibility of health care. Moreover, their connection with severe or deadly COVID-19 illness has further increased their societal relevance. Tools for early detection of these chronic diseases enable interventions to prevent high-impact complications, such as loss of sight and kidney failure. Similarly, prognostic tools for COVID-19 help stratify the population to prioritize protection and vaccination of high-risk groups, optimize medical resources and tests, and raise public awareness.

**METHODS:** We developed and validated state-of-the-art risk models for the presence of undiagnosed diabetes, hypertension, visual complications associated with diabetes and hypertension, and the risk of severe COVID-19 illness (if infected). The models were estimated using modern methods from the field of statistical learning (e.g., gradient boosting trees), and were trained on publicly available data containing health and socioeconomic information representative of the Mexican population. Lastly, we assembled a short integrated questionnaire and deployed a free online tool for massifying access to risk assessment.

**RESULTS:** Our results show substantial improvements in accuracy and algorithmic equity (balance of accuracy across population subgroups), compared to established benchmarks. In particular, the models: i) reached state-of-the-art sensitivity and specificity rates of 90% and 56% (0.83 AUC) for diabetes, 80% and 64% (0.79 AUC) for hypertension, 90% and 56% (0.84 AUC) for visual diminution as a complication, and 90% and 60% (0.84 AUC) for development of severe COVID disease; and ii) achieved substantially higher equity in sensitivity across gender, indigenous/non-indigenous, and regional populations. In addition, the most relevant features used by the models were in line with risk factors commonly identified by previous studies. Finally, the online platform was deployed and made accessible to the public on a massive scale.

**CONCLUSIONS:** The use of large databases representative of the Mexican population, coupled with modern statistical learning methods, allowed the development of risk models with state-of-the-art accuracy and equity for two of the most relevant chronic diseases, their eye complications, and COVID-19 severity. These tools can have a meaningful impact on democratizing early detection, enabling large-scale preventive strategies in low-resource health systems, increasing public awareness, and ultimately raising social well-being.

## 1. Introduction

The use of statistical models has contributed to the development of risk indices for the presence or future development of various diseases (e.g. diabetes, cancer, and cardiovascular disease) or the occurrence of disruptive health events (e.g. bone fractures and strokes)^1^. The deployment of these risk assessment models through online platforms seeks to aid with expanding access to disease prevention and health awareness and improving resource management in healthcare institutions.

The present work reports on the development of state-of-the-art models for estimating the risk of having diabetes and/or hypertension—two of the most common chronic diseases in Mexico and Latin America —as well as assessing some of the most relevant health risks associated with them: visual impairment and COVID-19 severity. In particular, leveraging large datasets representative of the Mexican population, we developed models that identify high-risk profiles for 1) diabetes and hypertension, 2) development of visual impairment as a complication of diabetes or hypertension (for patients with diabetes and/or hypertension), and 3) the development of severe COVID-19 disease leading to requiring intensive care or death (for COVID-19 infected patients). Overall, we show that the tool has substantial improvements in sensitivity and specificity on Mexican population, compared to the available benchmarks. Moreover, to maximize the accessibility and ease of use of the tool, we homogenized the variables in a single questionnaire and prioritized the use of information that is commonly known to an average person.

## 2. Diabetes and Hypertension

### Importance of Early Detection

According to the WHO, more than 500M people live with diabetes and 1.3Bn live with hypertension worldwide [28, 54]. Just for diabetes, its prevalence is predicted to continue growing at a 3% CAGR reaching 630 million people by 2045. Notably, both diseases cause a range of complications from which the most prevalent are neuropathy, kidney damage, visual impairment, heart attacks, and strokes. The progression of associated complications derives into a high economic and social impact due to early death and disability.

In Latin America and the Caribbean, the annual direct and indirect costs of diabetes and hypertension reached US $57.1 million in 2015 [10]. In Mexico, diabetes is the first cause of death and disability (DALYs) mainly due to the development of kidney failure, diabetic retinopathy, diabetic foot, and amputations [1, 43]. Moreover, the burden is expected to increase given that only 50% of the patients suffering from these two chronic diseases are aware of it [3, 32]. One of the crucial factors that influence the limited diagnosis rate is that approximately 60% of the population lacks social security and in consequence, has no access to preventive health services or adequate treatment [58]. Additionally, these conditions are associated with the progression and severity of other pathologies, such as Alzheimer’s disease and COVID-19.

Among many other benefits, an early diagnosis of these diseases can result in an 80% reduction of individual total annual healthcare expenditure, for example, from $12,500 to $2,500 in the USA [2]. This reduction is achieved through adequate monitoring and control of the disease that can prevent the development of major renal and ophthalmic complications. Additionally, early intervention strategies targeting habits, nutrition, and physical activity among people with a risk of developing diabetes and hypertension can reduce between 30% and 70% the probability of developing the disease and its complications in the long term [37, 23, 44].

### Current methods for risk estimation

There are numerous questionnaires to quantify individuals’ risk of having or developing diabetes and cardiovascular diseases [35, 31, 9, 39]. In 1993, the American Diabetes Association (ADA) published their first risk calculator to target undiagnosed diabetes based on decision tree predictive modeling. The model showed a sensitivity of 79% and specificity of 65% [36]. In 2009, Bang et al. [9] developed a screening tool based on multiple logistic regression for estimating the risk of having undiagnosed diabetes or prediabetes. The risk model was tested with information from persons of the U.S. population from different ethnic groups. This screening tool was adapted into a web-based risk calculator and is the current model employed by the ADA. Nowadays, another questionnaire widely used is the Finnish Diabetes Risk Score (FINDRISC) that calculates diabetes risk based on age, body mass index, waist circumference, physical activity, fruits and vegetable consumption, history of anti-hypertensive drug treatment, and history of high blood glucose [42]. Several adaptations of the FINDRISC have been proposed, such as the Latin American (LA-FINDRISC) and the Colombian versions (ColDRISC) [7, 49]. Despite its simplicity and wide usability, a recent evaluation of FINDRISC reported a sensitivity of 70% and specificity of 67% [11].

Specifically for hypertension risk assessment, the available models are more scarce and inaccurate, it is more common to find risk calculators for acute cardiovascular diseases such as myocardial infarction, angina pectoris, or stroke. One of the few risk models for hypertension was developed by Kshirsagar et al. [39], who created a scoring algorithm to determine the risk of developing hypertension in a time-lapse of 3, 6, and up to 9 years. This risk model was fitted to a predominantly white population (83%) of middle age and older adults.

Overall, systematic reviews have found varying results. A recent review published in 2018 evaluated 73 different risk calculators for CVD and found a wide variation among estimates for the same patient. Furthermore, the authors concluded that most calculators have deficient communication strategies that translate to low actionability [14]. Similar systematic analyses of diabetes risk calculators reported a lack of statistical robustness, risk overestimations, and limited implementability due to the requirement of specialized lab tests [50, 5, 17]. However, the increasing availability of large databases with clinical and sociodemographic information of local populations, as well as the availability of modern statistical learning methods, pose an opportunity to raise accuracy and adaptability to local populations [15, 38, 57].

### 2.1. Methodology

#### Data

For the training and validation of the diabetes and hypertension risk models, we used the database of the Mexican National Health and Nutrition Survey 2018 (ENSANUT 2018). The ENSANUT survey is the result of a joint effort of the Mexican Health Secretary, the National Institute of Public Health (INSP), and the National Institute of Statistics and Geography (INEGI). The main goal of the ENSANUT is to collect detailed information on the health status and nutritional conditions of the pediatric and adult Mexican population. The ENSANUT is carefully designed to be representative of the Mexican population at the national and state levels, with a thorough territorial and sociodemographic sampling strategy. The 2018 edition collected information on 50,000 households distributed among the 32 states. In total, the ENSANUT 2018 contains information of more than 60,000 people from 4 age groups: pre-school children (0-4 years old), school-age children (5-9 years old), teenagers (10-19 years old), and adults (20 and more) [60].

For the present project, the data from the 43,071 adults surveyed was used. We divided the variables into the following categories: basic information, sociodemographics, general health information, chronic diseases, health habits, physical functionality, physical activity, anthropometric measures, and diet. We further divided all categories into subcategories.

To account for the undiagnosed population, we used the data of respondents for which a biomarker was available. For diabetes, we considered respondents with a glycated hemoglobin (HbA1c) test (*n* = 12, 919) and for hypertension those with blood pressure measurements (*n* = 17, 474). We classified people as diabetic when they presented a medical diagnosis or a measured HbA1c level of 6.4% or greater [4]. For hypertension, we considered individuals with a medical diagnosis or a measurement of either more than 140 mmHg systolic pressure, or more than 90 mmHg diastolic pressure [46].

#### Selection of Variables

To find an optimal set of predictive variables we first identified questions that were appropriate to include and could be answered by most respondents without the help of a medical professional and without any biochemical measurement. Afterwards, we employed a forward-stepwise regression that added a new subcategory of questions at each step [33]. The criterion for the procedure was to maximize the out-of-sample area under the receiver operating characteristic curve (AUC) of a Lasso logistic model trained using 5 fold cross-validation [27, 61, 34]. Thereafter, the resulting set of questions was reduced via a backward-stepwise regression removing one variable at each step, following the same criterion as before [34]. Lastly, few manual adjustments were made to produce a common set of variables for both models without sacrificing accuracy.

#### Model and Validation

Several statistical algorithms were compared, with the best results yielded by *gradient boosting classifiers*. To avoid overfitting the following regularization hyperparameters were introduced: number of classifiers, learning rate, subsample, the maximum number of available features to a single classifier, and the minimum number of samples per leaf [29, 34]. The hyperparameters were tuned using grid search with cross-validation [40].

All the reported evaluation metrics, such as the area under the receiver operating characteristic curve (AUC), sensitivity, specificity, and the disparity between subgroups were calculated using **out-of-sample validation** scores (based on 10-fold cross-validation [34]), and confidence intervals for such metrics were computed using bootstrap sampling [25].

#### Importance of Features and Equity across Subgroups

##### Importance of features

Feature importance was measured by sensitivity analysis based on permuted values [53, 6, 16]. In particular, for each variable, the method randomly permutes the values among all observations in the dataset and compares the risk scores before and after the permutation. Variables for which the risk scores before and after permutation differ the most are the most sensitive and important for the model.

##### Equity across subgroups

To measure the disparity or equity of model performance across subgroups, we first calculate the sensitivity of the models for each population subgroup (setting a common discrimination threshold corresponding to 55% specificity). We then measure disparity in model performance as the average difference between the model sensitivity (*S*_*g*_) on each group *g* and the sensitivity (*S*) of the model on the population as a whole: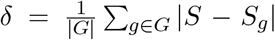. 95% confidence intervals for each sensitivity were computed using bootstrap sampling [25].

The relevant dimensions for population segmentation used were: sex (male/female), origin (indigenous/non-indigenous)^2^, and geographic region.

#### Benchmark Methodologies

##### Diabetes

The diabetes model was benchmarked against the aforementioned risk calculator developed by ADA [9] and the LA-FINDRISC risk score [7].

To perform a thorough evaluation of the ADA risk calculator on a Mexican population, we developed an automated computer program that transformed, fed, and evaluated a large representative sample of the Mexican population from ENSANUT (*n* = 9, 794) to the web-based calculator made available by ADA^3^. The output of the web-based ADA calculator was recorded for each ENSANUT observation and compared against the ENSANUT ground-truth to assess its accuracy on the Mexican population.

For the LA-FINDRISC risk score, we replicated the algorithm following the specifications given by the authors and evaluated its performance with the data of 9,318 respondents of the ENSANUT for which all required variables were available.

##### Hypertension

The algorithm proposed by Kshirsagar et al. [39] was implemented as a benchmark for the hypertension model, and its performance was evaluated on a representative sample of 11,367 respondents from ENSANUT. Because the purpose of the present work is to build and evaluate risk assessment tools that do not require any biological measurement, we implemented a version of the Kshirsagar et al. algorithm that does not include blood pressure as an input variable.

### 2.2. Results

#### Diabetes

##### Accuracy

Substantial accuracy improvements were achieved by the model compared with the benchmarks (Figure 2). Overall, the model achieved an AUC of 0.83 in out-of-sample scores, in comparison with an 0.74 AUC of the ADA benchmark and 0.75 AUC of the LA-FINDRISC benchmark (a 12% and 11% relative improvement respectively). In particular, Figure 2 shows that the model can increase sensitivity from 80% to 90% (a 13% relative improvement) compared to the benchmarks while maintaining specificity constant at 56%. Similarly, it shows that the model can increase specificity from 56% to 70% (a 25% relative improvement) compared to the benchmarks while maintaining sensitivity constant at 80%.

**Figure 1:**
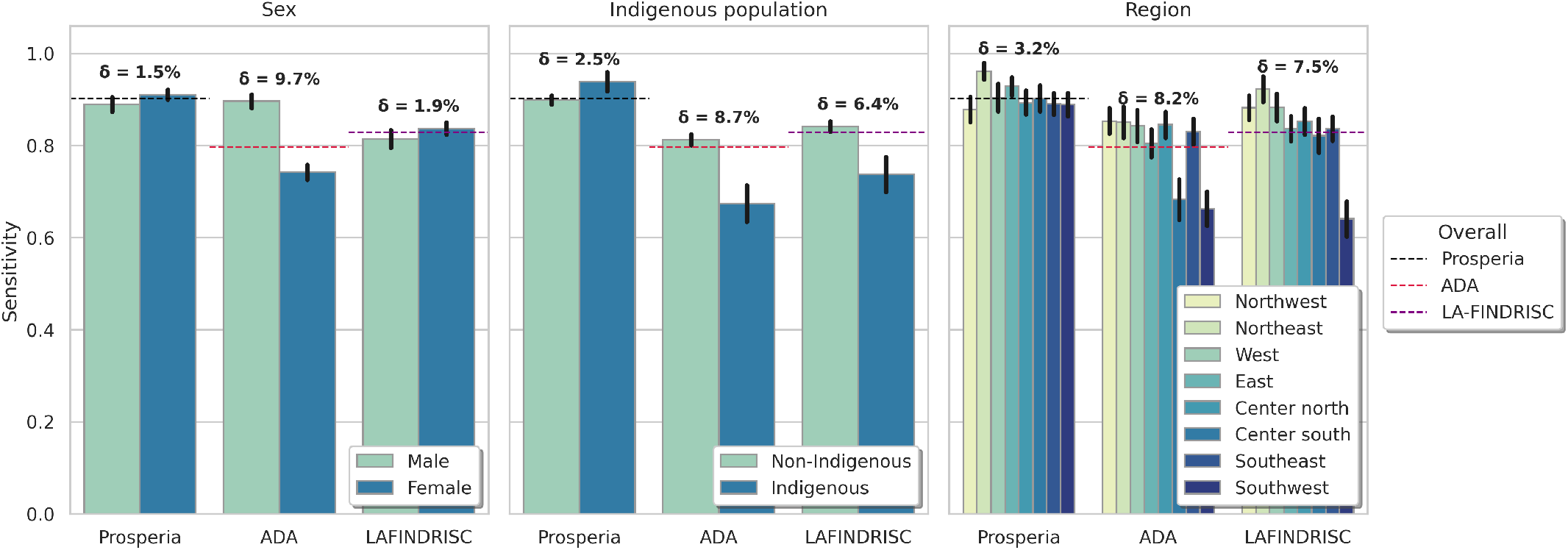
Equity across subgroups for the diabetes model and benchmark.

**Figure 2:**
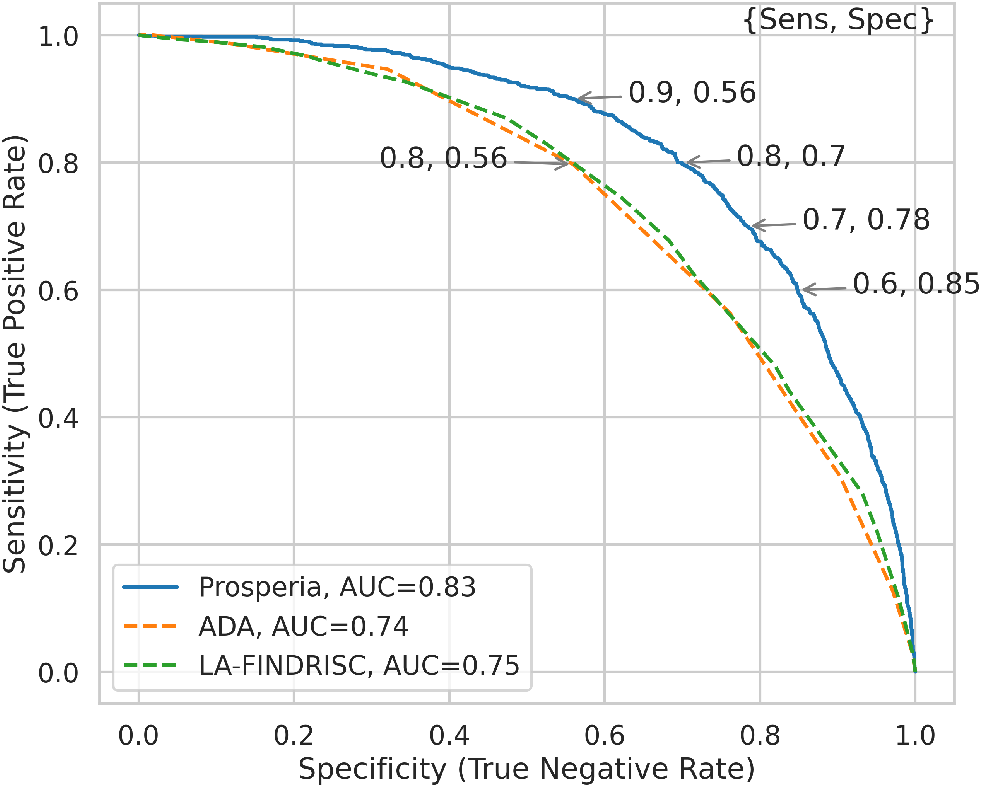
ROC curves of the diabetes model and benchmarks.

##### Importance of features

The variable with the largest impact on the diabetes risk index was found to be the *age*, followed by *family history of diabetes, recent change of weight*, and current *weight* (Figure 3). Comorbidities associated with metabolic syndrome such as *obesity, high levels of cholesterol and triglycerides*, and *hypertension* follow in contribution. Demographic and socioeconomic variables, including *education level* and *region of residence*, as well as *physical activity*, also showed considerable importance.

**Figure 3:**
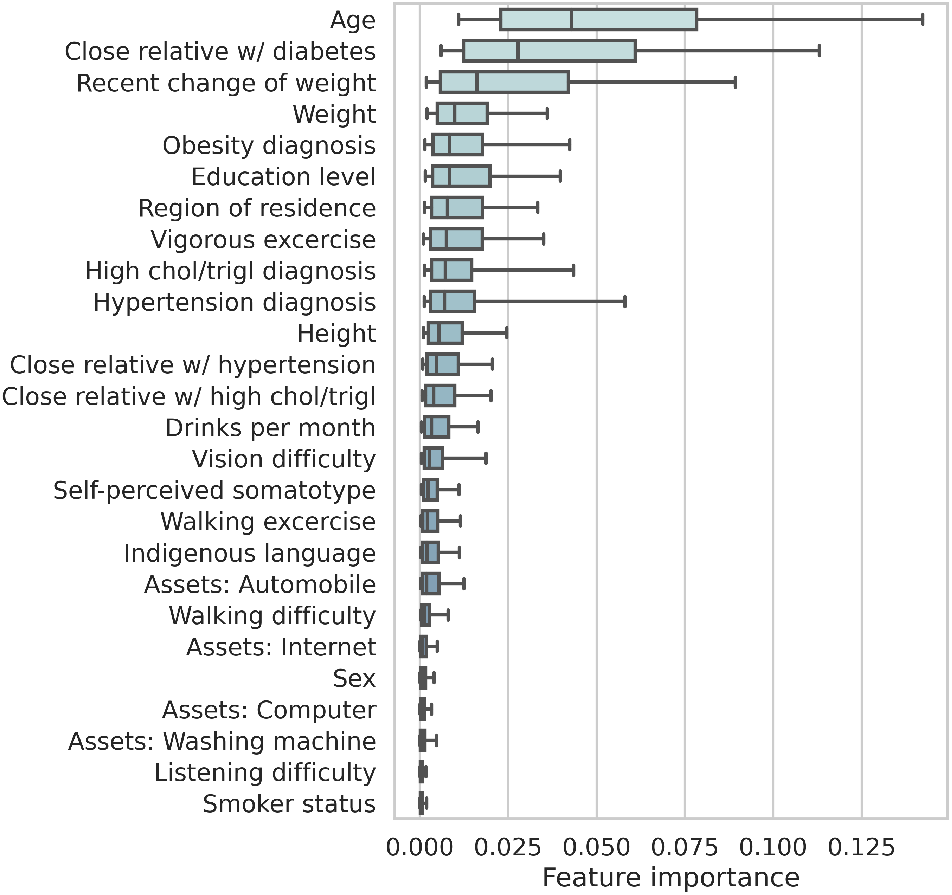
Importance of features for the diabetes model.

##### Equity across subgroups

The ADA benchmark shows considerable levels of inequity (Figure 1); with a disparity of 9.7% across sex groups, 8.7% between indigenous and non-indigenous population, and 8.2% across regions. Specifically, the ADA benchmark shows a lower sensitivity for the female and indigenous people, the latter dropping below 70%. The LA-FINDRISC benchmark showed lower levels of inequity with a disparity of 1.9% across sex groups, 6.4% between indigenous and non-indigenous population, and 7.5% across regions. In comparison, our model performed well across all subgroups, with low disparity at 1.5%, 2.5%, and 3.2% for each respective division.

##### Hypertension

###### Accuracy

Substantial accuracy improvements were achieved by the model compared with the Kshirsagar et al. benchmark (Figure 5). Overall, the model achieved an AUC of 0.79 in out-of-sample scores, in comparison with an 0.67 AUC of the benchmark (an 18% relative improvement). In particular, Figure 5 shows that the model can increase sensitivity from 73% to 85% (a 16% relative improvement) compared to the benchmark while maintaining specificity constant at 50%. Similarly, it shows that the model can increase specificity from 50% to 70% (a 40% relative improvement) compared to the benchmark while maintaining sensitivity constant at 73%.

**Figure 4:**
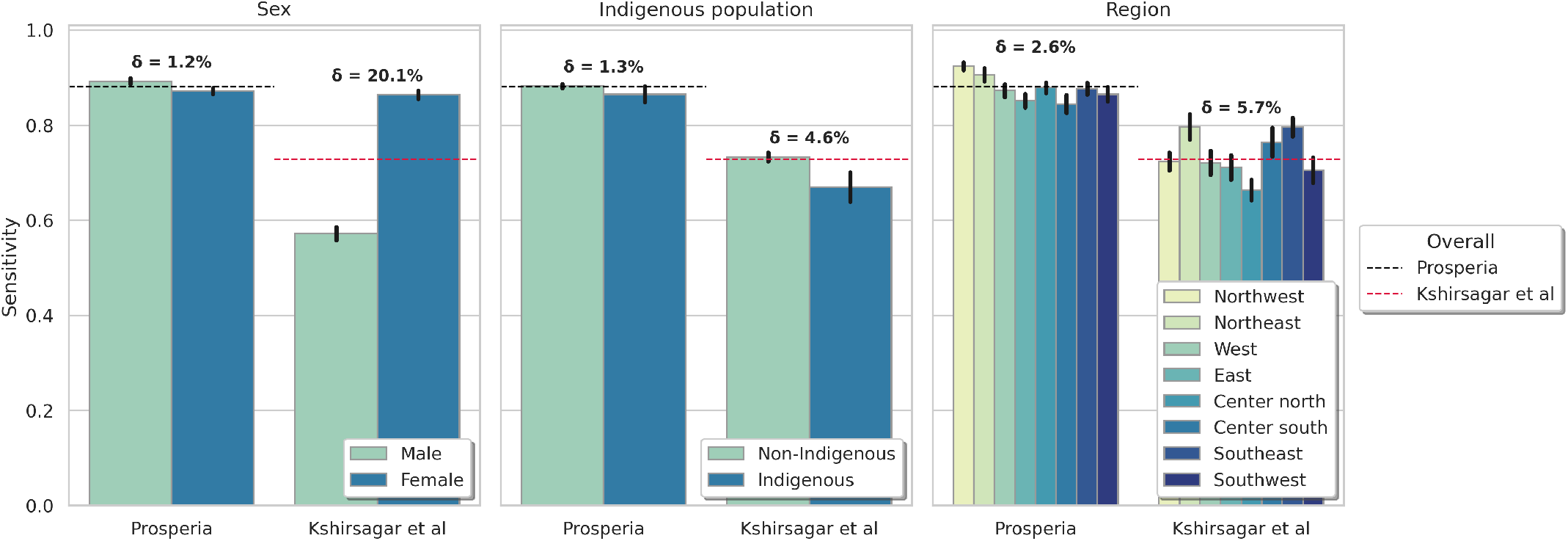
Equity across subgroups for the hypertension model and benchmark.

**Figure 5:**
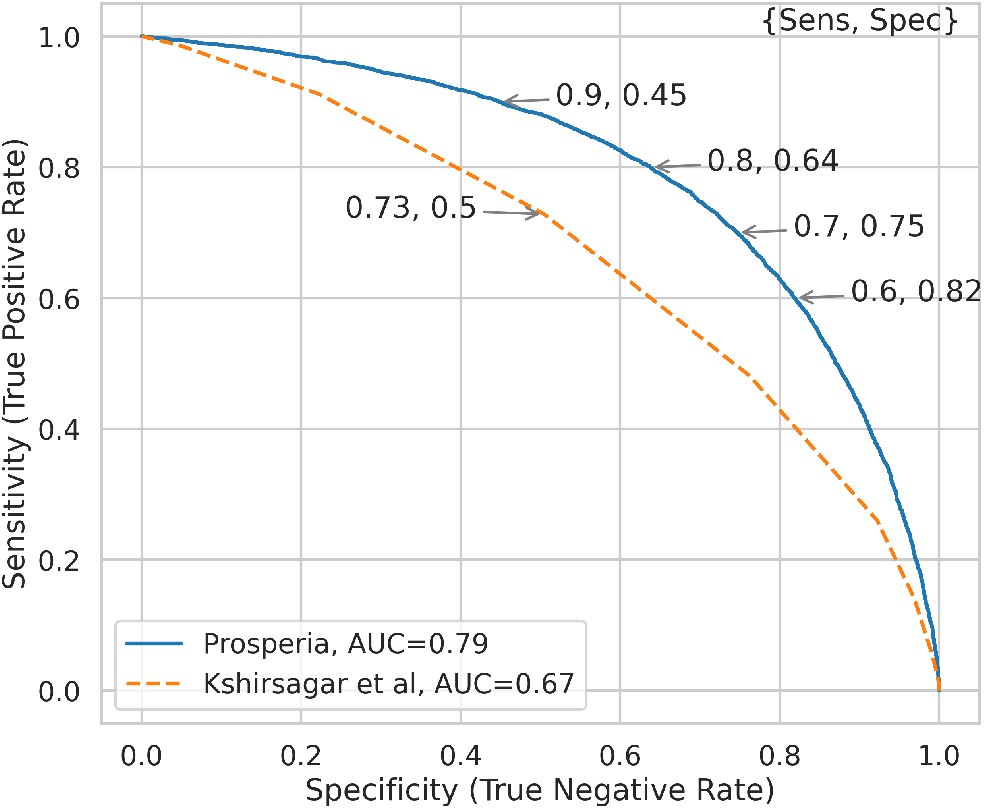
ROC curves of the hypertension model and benchmark.

###### Importance of features

The variable with the largest impact on the hypertension risk index was found to be the *age* (Figure 6). Other highly relevant variables include *weight* and *family history of hypertension*, followed by demographic and socioeconomic variables as well as comorbidities such as *diabetes, high levels of cholesterol and triglycerides*, and *obesity*.

**Figure 6:**
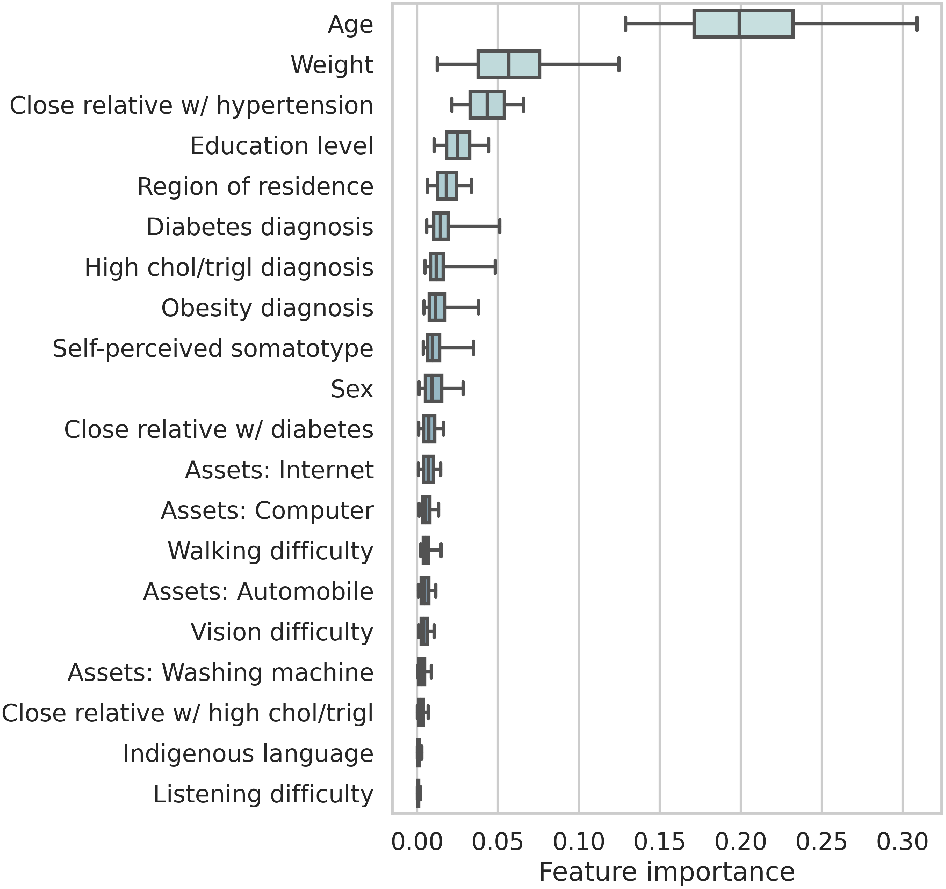
Importance of features for the hypertension model.

###### Equity across subgroups

Our model performed equally well in all subdivisions with a disparity of 1.2% between sex groups, 1.3% between indigenous and non-indigenous population, and 3.6% across regions of residence. In comparison, Figure 4 shows that the benchmark has a rather substantial gap in performance between sex groups, with the sensitivity dropping below 60% for the male population; and a more moderate performance gap between indigenous and non-indigenous groups, with higher sensitivity for the non-indigenous population.

## 3. Diabetic and Hypertensive Retinopathy

### Importance of Early Detection

The role of hypertension and diabetes in vision loss represents both a challenge and an opportunity to improve patients’ life quality and outcomes [18]. Persistent high blood glucose levels, as well as high blood pressure, causes blood vessel abnormalities in the retina—diabetic or hypertensive retinopathy—characterized by thickening or blockage of the arteries, swelling, bleeding, and in severe cases, macular edema and/or retina detachment [65, 12].

The prevalence of diabetic retinopathy (DR) is of significant importance globally. One in three people with diabetes develops some degree of retinopathy over their lifetime [66], making it the leading cause of vision loss in working-age adults. This complication and its increasing prevalence threatens particularly, the public health institutions in developing countries like Mexico, where it represents the first cause of irreversible blindness in the country, causing great economic impact due to direct and indirect costs, including loss of productivity and payment of disability insurances.

However, a key opportunity for reducing the social and economic impact of vision-threatening retinopathy is to target the disease at early stages (i.e. mild and moderate retinopathy) when vision loss is still preventable. However, considering the absence of salient symptoms at early stages [52], this strategy requires the development of awareness campaigns and accurate early-detection tools.

### Current methods for Risk Estimation

Several questionnaires exist to quantify individuals’ risk of having or developing sight-threatening diabetic retinopathy [51, 62, 26], however, we couldn’t identify any available risk assessment tool for hypertensive retinopathy. One screening tool with substantial usage in Europe is the model developed by Aspelund et al. which is used to estimate the risk of developing sight-threatening retinopathy in one year after an eye revision, as well as giving a time-lapse recommendation for the following examination [8]. The algorithm was fitted and tested in a Nordic population (Iceland and Denmark) with supporting validation on several European countries. In Mexico, Mendoza-Herrera et al. 2017 developed a screening tool for diabetic retinopathy, which consists of two self-declared variables (age and physical activity) and two measurements (blood pressure and capillary glucose level) [47].

### 3.1. Methodology

#### Data

To develop the models for diabetes-related visual diminution and hypertensive retinopathy, we employed the subsets of diagnosed diabetic and hypertensive patients from the ENSANUT 2018 data set. These components of the survey include several variables related to the treatment and progression of each disease [60].

#### Selection of Variables

Following the objective of creating a single questionnaire for all risk calculators, we started with the set of variables from the diabetes and hypertension models. Then, for the model of diabetes-related visual diminutions, we created a new set of variables by adding several questions regarding diabetes progression and treatment that we determined were appropriate to include and could be answered by most respondents without the help of a medical professional, and without any biochemical measurement. Similarly, we constructed a set of variables for the hypertensive retinopathy model. Thereafter, each resulting set of questions was reduced via a backward-stepwise regression removing one variable at each step following the criterion of maximizing the out-of-sample area under the receiver operating characteristic curve (AUC) of a Lasso logistic model trained using 5-fold cross-validation [27, 61, 34]. Lastly, few manual adjustments were made to produce a common set of variables for both models without sacrificing accuracy.

#### Model and Validation

Several statistical algorithms were compared, with the best results yielded by *gradient boosting classifiers*. To avoid overfitting the following regularization hyperparameters were introduced: number of classifiers, learning rate, subsample, the maximum number of available features to a single classifier, and the minimum number of samples per leaf [29, 34]. The hyperparameters were tuned using grid search with cross-validation [40].

All the reported evaluation metrics, such as the area under the receiver operating characteristic curve (AUC), sensitivity, specificity, and the disparity between subgroups were calculated using **out-of-sample validation** scores (based on 10-fold cross-validation [34]), and confidence intervals for such metrics were computed using bootstrap sampling [25].

#### Ensemble Model

To create a more general screening tool for retinal damage caused by metabolic disorders, we created a model for both diabetes-related visual diminution and hypertensive retinopathy. Towards it, we implemented a simple ensemble model that outputs the highest risk score of the two models.^4^ For evaluation, we applied the ensemble model out-of-sample on the ENSANUT data and compared it against its ground-truth.

#### Importance of Features and Equity across Subgroups

##### Importance of features

Feature importance was measured by sensitivity analysis based on permuted values [53, 6, 16]. In particular, for each variable, the method randomly permutes the values among all observations in the dataset and compares the risk scores before and after the permutation. Variables for which the risk scores before and after permutation differ the most are the most sensitive and important for the model.

##### Equity across subgroups

To measure the disparity or equity of model performance across subgroups, we first calculate the sensitivity of the models for each population subgroup (setting a common discrimination threshold corresponding to 60% specificity). We then measure disparity in model performance as the average difference between the model sensitivity (*S*_*g*_) on each group *g* and the sensitivity (*S*) of the model on the population as a whole: 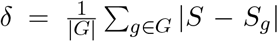. 95% confidence intervals for each sensitivity were computed using bootstrap sampling [25].

The relevant dimensions for population segmentation used were: sex (male/female), origin (indigenous/non-indigenous)^5^, and geographic region.

##### Benchmark Methodologies

###### Diabetes-related visual diminution

We considered two risk models as benchmarks: Mendoza-Herrera’s model based on data from a Mexican population [47] and Aspelund’s model fitted and tested with a Nordic population [8].

Mendoza-Herrera et al., employed a probit model to determine the risk of undiagnosed diabetic retinopathy [47]. We replicated their model following the specifications given by the authors and evaluated its performance with the data of 1348 diabetic respondents of the ENSANUT for which all required variables were available.

The screening tool developed by Aspelund et al., employs a Weibull proportional hazards model. The main objective of this tool is estimating the risk of developing sight-threatening retinopathy in the following 12 months after an examination [8]; however, the same statistical model can calculate the risk of developing this condition at a given time of diabetes progression [59]. We implemented this second version of the model, following the specifications of the authors for the parameters of the model, and evaluated its performance with the data of 1348 diabetic respondents of the ENSANUT for which all required variables were available.

###### Hypertensive retinopathy

No benchmark was found for hypertensive retinopathy.

### 3.2. Results

#### Diabetes-Related Visual Diminution

##### Accuracy

Substantial accuracy improvements were achieved by the model compared with the benchmarks (Figure 8). Overall, the model achieved an AUC of 0.76 in out-of-sample scores, in comparison with an 0.61 AUC of the Mendoza-Herrera et al. benchmark and 0.62 AUC of the Aspelund et al. benchmark (a 25% and 23% relative improvement respectively). In particular, Figure 8 shows that the model can increase sensitivity from 68% to 85% (a 25% relative improvement) compared to the benchmarks while maintaining specificity constant at 48%. Similarly, it shows that the model can increase specificity from 48% to 67% (a 40% improvement) compared to the benchmarks while maintaining sensitivity constant at 68%.

**Figure 7:**
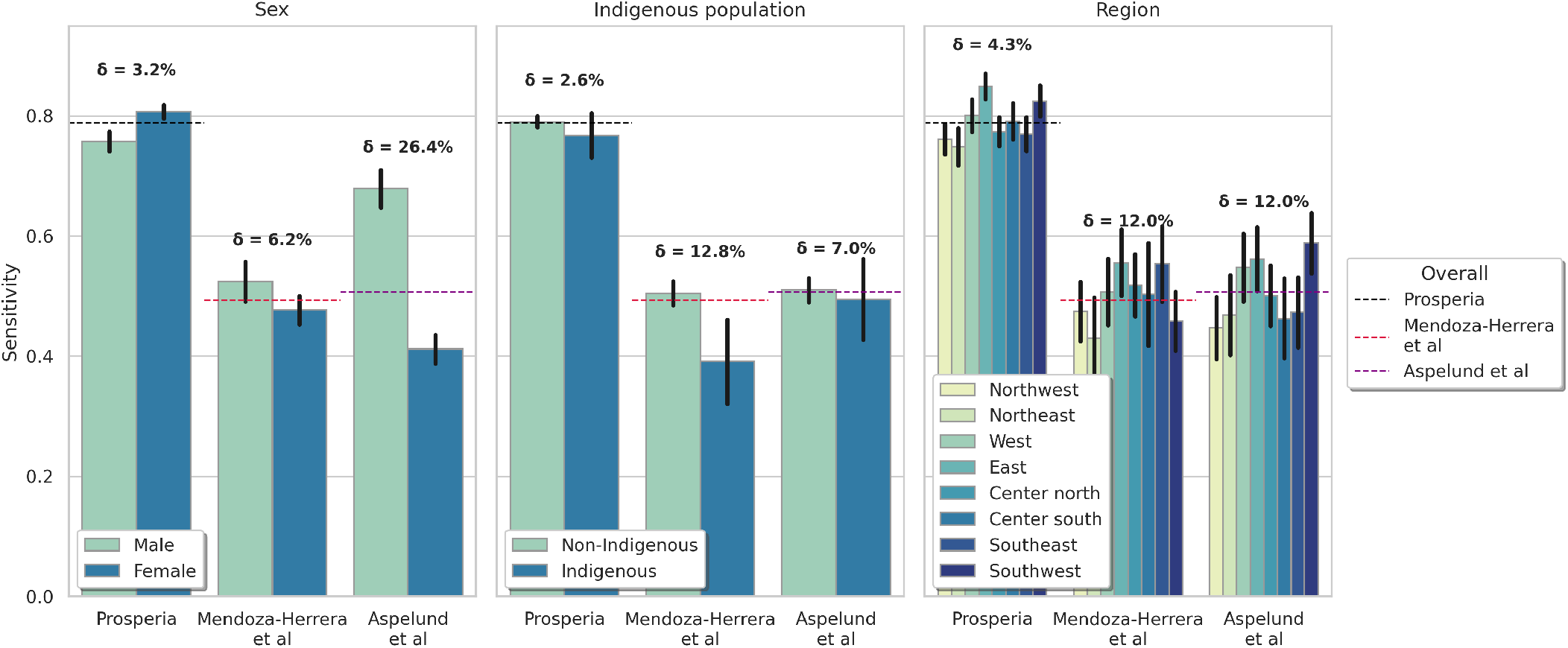
Equity across subgroups for the diabetes-related vision diminution model and benchmarks.

**Figure 8:**
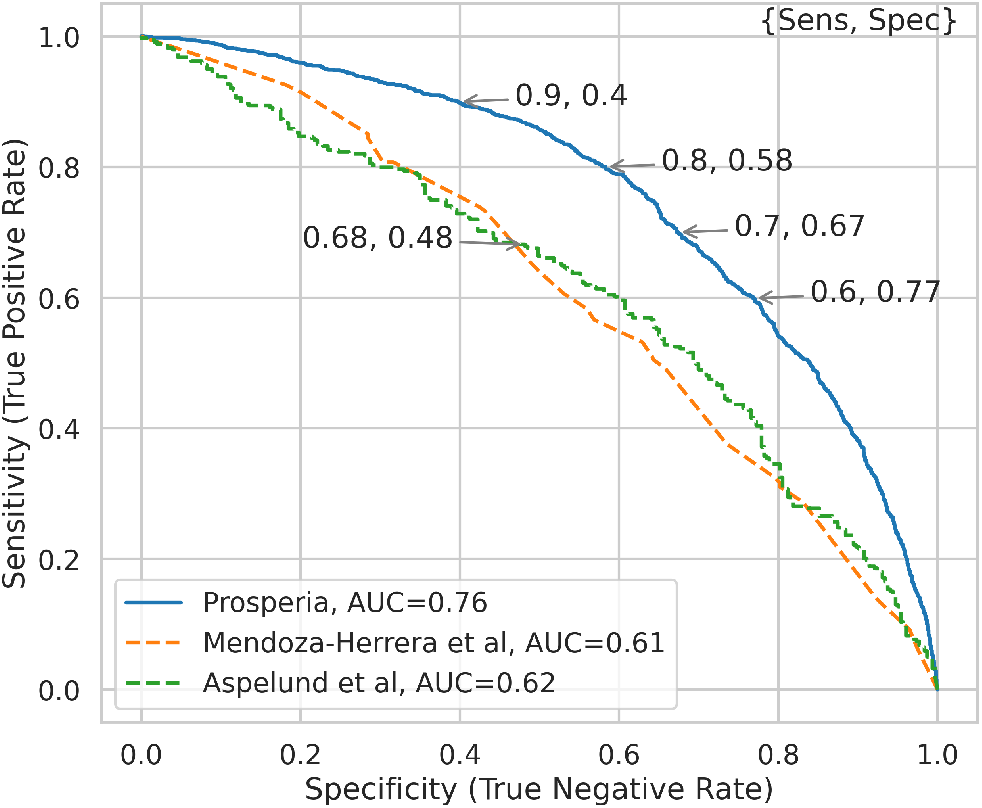
ROC curves of the diabetes-related visual diminution model and benchmarks.

##### Importance of features

The variable with the largest impact on the risk index was found to be the perceived *difficulty to see* (Figure 9). Following in importance are variables related to the treatment and progression of diabetes such as *diabetes evolution time, medication, complementary treatment*, and *time since the last medical visit*. Other highly relevant variables are *age, depression, trouble sleeping*, and *urinary tract infections*.

**Figure 9:**
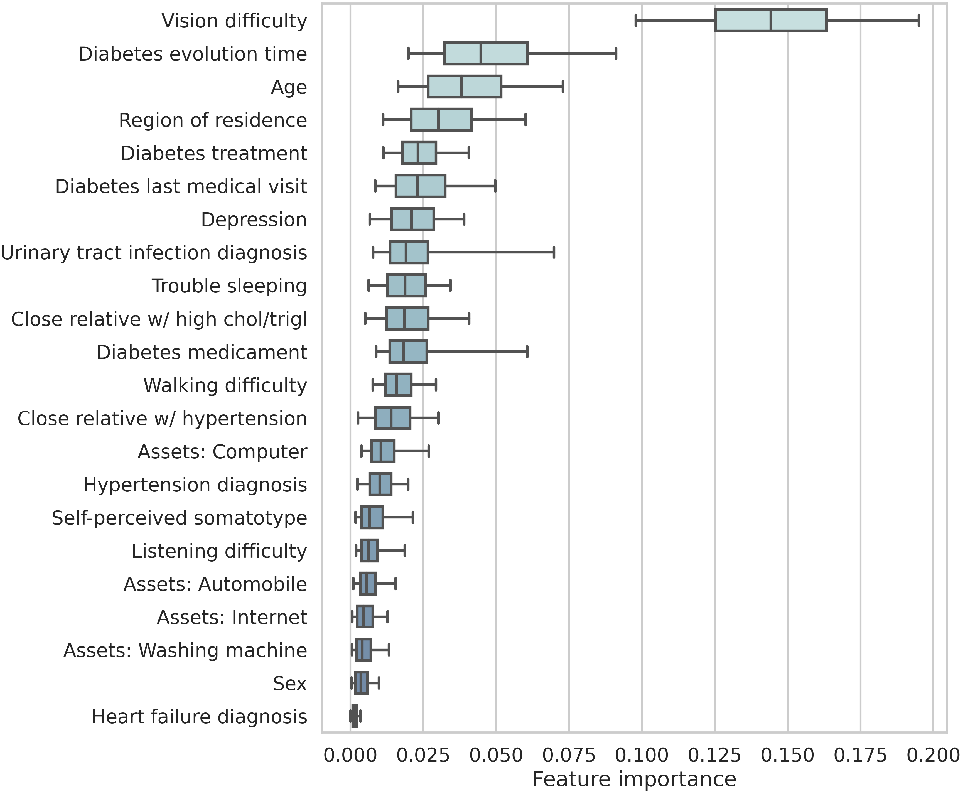
Importance of features for the diabetes-related visual diminution model.

##### Equity across subgroups

The benchmark from Aspelund et al. shows a very large difference between the sensitivity of the model in the male and female populations, favoring the first (Figure 7). The benchmark from Mendoza-Herrera et al. shows a considerable disparity between indigenous and non-indigenous populations, favoring the latter. Both benchmarks show a large disparity across regions. In comparison, our model performed similarly well in all subdivisions with a disparity of 3.2% between sex groups, 2.6% between indigenous and non-indigenous population, and 4.3% across regions of residence.

##### Hypertensive Retinopathy

###### Accuracy

The model performed reasonably well, achieving an AUC of 0.76 in out-of-sample scores. Figure 10 indicates the sensitivity and specificity of the model at different discrimination thresholds. As mentioned in the subsection 3.1, there were no relevant benchmarks found for hypertensive retinopathy risk calculators.

**Figure 10:**
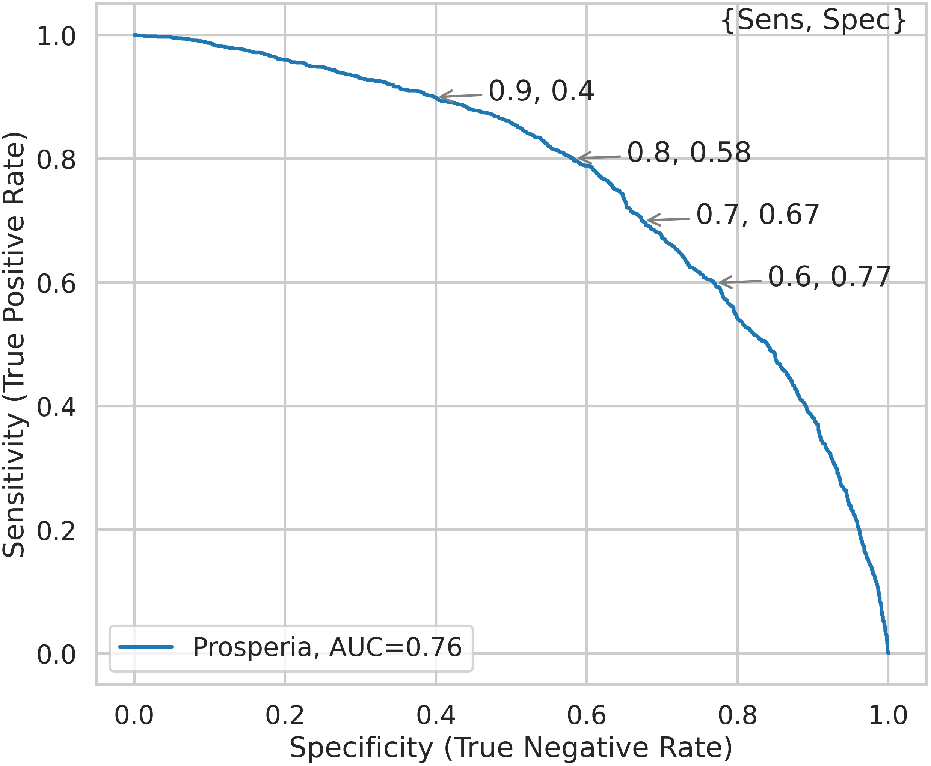
ROC curve of the hypertensive retinopathy model.

###### Importance of features

The variable with the largest impact on the risk index was found to be the perceived *difficulty to see*, followed by *depression*, variables related to the treatment of hypertension, and *urinary tract infections* (Figure 11). Other highly relevant variables include *age, weight*, having *difficulty sleeping*, and *region of residence*.

**Figure 11:**
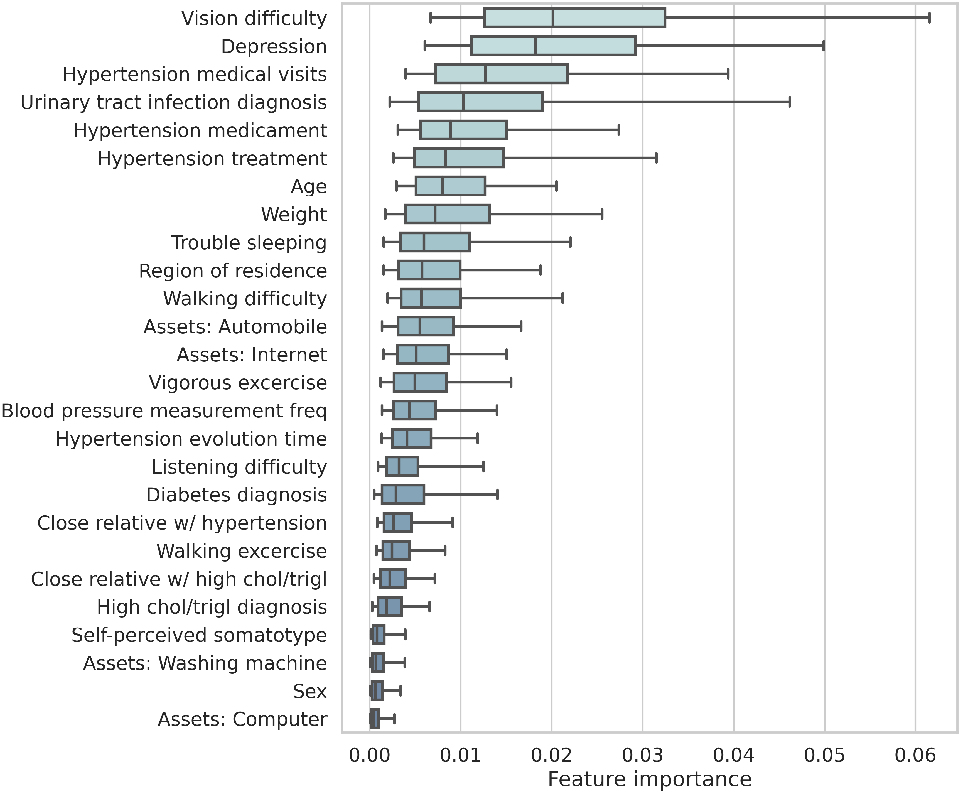
Importance of features for the hypertensive retinopathy model.

###### Equity across subgroups

Our model showed considerable disparities across subgroups: 10.4% between sex groups, 9.6% between indigenous and non-indigenous population, and 8.0% across regions of residence. Figure 12 shows that the model has a higher sensitivity in the female and indigenous populations. However, as shown below, the visual diminution ensemble model was able to mitigate them.

**Figure 12:**
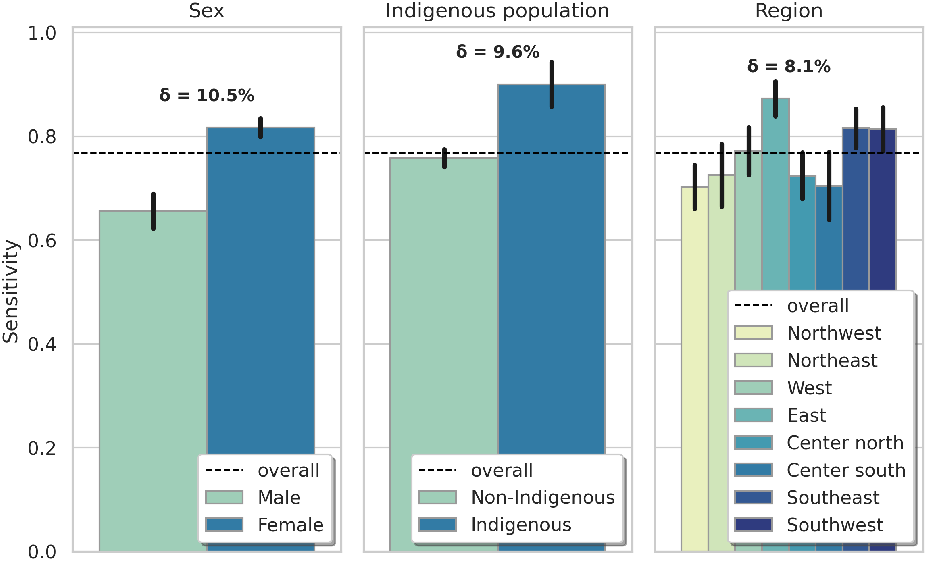
Equity across subgroups for the hypertensive retinopathy model.

##### Visual Diminution Ensemble Model

###### Accuracy

A substantial improvement in precision was achieved with the visual diminution ensemble model, in comparison with each of its independent components: diabetic and hypertensive vision diminution. Overall, the model achieved an AUC of 0.84, in comparison with a 0.76 AUC for both independent models (an 11% relative improvement). In particular, Figure 13 shows that the model can increase sensitivity from 80% to 90% (a 13% relative improvement) compared to the independent models while maintaining specificity around 55%. Similarly, it shows that the model can increase specificity from 55% to 72% (a 31% relative improvement) compared to the independent models while maintaining sensitivity constant at 80%.

**Figure 13:**
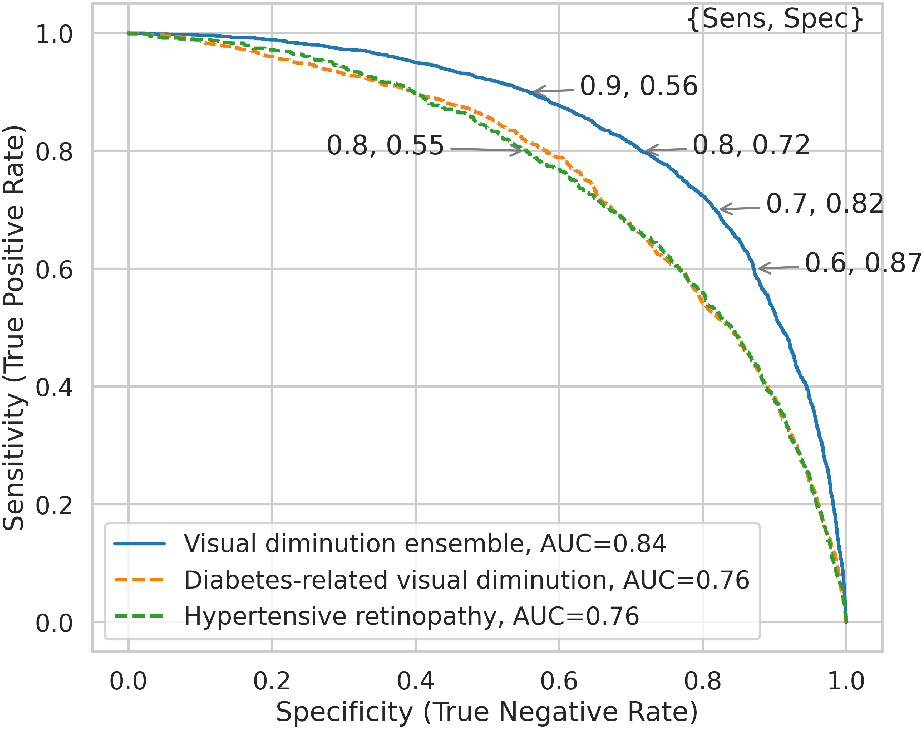
ROC curves of the visual diminution ensemble model and components.

###### Equity across subgroups

The ensemble of visual diminution models also greatly improved the equity across subgroups. Figure 14 shows that the ensemble model achieves very low levels of disparity, at around 2% for all segmentation variables (sex, indigenous/non-indigenous, and regions), much lower than disparity levels found for the independent models.

**Figure 14:**
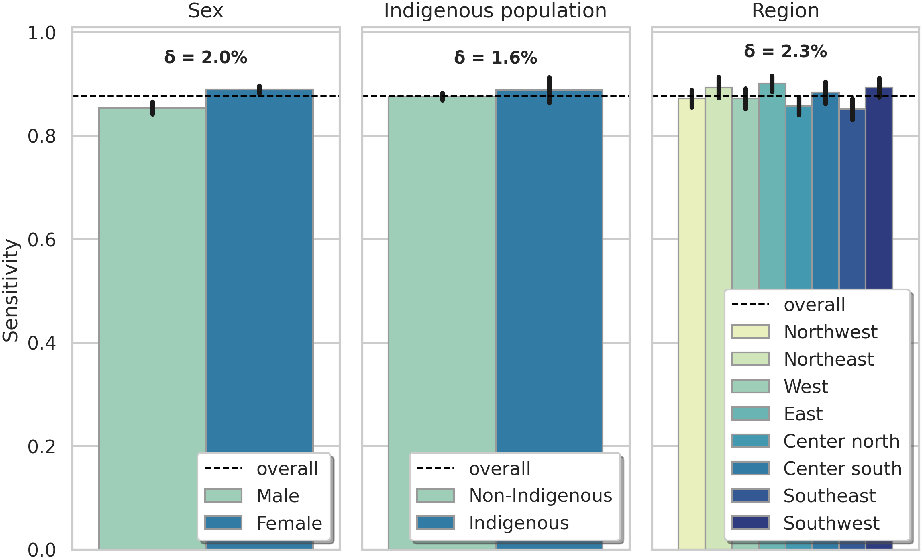
Equity across subgroups for the visual diminution ensemble model.

## 4. COVID-19 Severity

### Relevance of COVID-19 Severity Risk Stratification

Several studies have identified age, weight, sex, and comorbidities such as diabetes, hypertension, and asthma to be relevant risk factors associated with more severe cases of COVID-19 [64, 13]. Although most low and middle-income countries have a younger population, they show a higher infection fatality rate (IFR) than high-income countries; attributed to deficiencies in the quality and accessibility of health care. Estimations show that the IFR of similar diseases to COVID-19 doubles in upper-middle-income countries, nearly triples in lower-middle-income countries, and increases by a factor of 3.7 in low-income countries [30]. Additionally, most diabetic and hypertensive patients live in low and middle-income countries; with roughly 80% for diabetes and 65% for hypertension [32, 28, 54]. Mexico for example is among the ten countries with the highest mortality rates from COVID-19 (∼8.7%) [56]. There, 20% of patients with confirmed COVID-19 diagnosis suffer from diabetes or hypertension, and, of all fatal cases, 50% suffered from at least one of these comorbidities [19, 22].

It is in low and middle-income contexts that prognostic tools for COVID-19 severity can be most useful. There, governments mostly depend on non-pharmaceutical policies to avoid hospital saturation. Thus, surveillance and testing must be improved to reduce the spread of infection and to tailor efficient interventions [63]. Accurate prognostic tools for severe illness of COVID-19, accounting for undiagnosed comorbidities, can improve stratification of the population to prioritize protection and vaccination of high-risk groups, optimize medical resources and tests, as well as to raise public awareness.

### Current methods for Risk Estimation

Several institutions from different countries have implemented tools for evaluating the risk and severity of COVID-19 [41, 30, 64, 45]. However, most calculators assume user knowledge about their potential comorbidities like diabetes and hypertension, key risk factors, disregarding the high rate of underdiagnosis in developing countries [32, 28]. In Mexico, the Social Security Institute (IMSS) developed a calculator to estimate the risk of COVID-19 complications based on basic health information and present comorbidities [21]. This prognostic tool was adapted into a web-based risk calculator and is available to the public.

### 4.1. Methodology

#### Data

For the training and validation of the COVID-19 complications risk model, we used the data on COVID-19 cases from the Mexican General Directorate of Epidemiology (DGE) [24]. The dataset consists of the results of all the COVID-19 tests applied in Mexico—amounting to more than 1.6 million tests and 730,317 positive cases—relevant information about each tested person, and the outcome of the positive cases (whether or not the patient required hospitalization, intensive care, intubation, or died).

We only considered patients with a positive diagnosis of COVID-19 that were older than 20 years and excluded active ambulatory patients (with less than 15 days since symptoms onset and no hospitalization nor death) [55]. We classified as severe all the cases of patients that developed complications that resulted in death or that required intensive care.

#### Selection of Variables

Following the objective of creating a single questionnaire for all risk calculators, we started with the set of variables used by the diabetes and hypertension models. This set already accounted for most risk factors associated with severe cases of COVID-19: age, sex, diabetes, hypertension, obesity, and smoking [13, 64]. We fitted one model to the data using only variables available in the set and a second one with added risk factors such as the presence of chronic obstructive pulmonary disease, chronic renal disease, and immunosuppression. The added variables yielded no significant improvements in AUC compared with the basic set of variables, and thus we did not include them in the final version of the model.

#### Model and Validation

Several statistical algorithms were compared, with the best results yielded by *gradient boosting classifiers*. To avoid overfitting the following regularization hyperparameters were introduced: number of classifiers, learning rate, subsample, and the minimum number of samples per leaf [29, 34]. The hyperparameters were tuned using grid search with cross-validation [40].

All the reported evaluation metrics, such as the area under the receiver operating characteristic curve (AUC), sensitivity, specificity, and the disparity between subgroups were calculated using **out-of-sample validation** scores (based on 10-fold cross-validation [34]), and confidence intervals for such metrics were computed using bootstrap sampling [25].

#### Importance of Features and Equity across Subgroups

##### Importance of features

Feature importance was measured by sensitivity analysis based on permuted values [53, 6, 16]. In particular, for each variable, the method randomly permutes the values among all observations in the dataset and compares the risk scores before and after the permutation. Variables for which the risk scores before and after permutation differ the most are the most sensitive and important for the model.

##### Equity across subgroups

To measure the disparity or equity of model performance across subgroups, we first calculate the sensitivity of the models for each population subgroup (setting a common discrimination threshold corresponding to 60% specificity). We then measure disparity in model performance as the average difference between the model sensitivity (*S*_*g*_) on each group *g* and the sensitivity (*S*) of the model on the population as a whole: 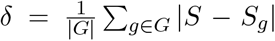. 95% confidence intervals for each sensitivity were computed using bootstrap sampling [25].

The relevant dimensions for population segmentation used were: sex (male/female), origin (indigenous/non-indigenous)^6^, and geographic region.

##### Benchmark Methodologies

We used the prognostic tool developed by the IMSS as a benchmark for the COVID-19 severity model. To evaluate its performance, we developed an automated computer program that transformed, fed, and evaluated the data from the DGE data set (*n* = 179, 889) to the web-based calculator^7^. The output of the web-based IMSS calculator was recorded for each DGE observation and compared against the DGE ground-truth to assess its accuracy.

### 4.2. Results

#### Accuracy

Both our model and the IMSS benchmark achieved high accuracy scores (Figure 16). However, a considerable improvement in sensitivity was still possible: the model achieved an AUC of 0.84 in out-of-sample scores, in comparison with an 0.80 AUC of the benchmark (a 5% relative improvement). In particular, Figure 16 shows that the model can increase sensitivity from 85% to 90% (a 6% relative improvement) compared to the IMSS benchmark while maintaining specificity at around 60%. Similarly, it shows that the model can increase specificity from 61% to 68% (an 11% relative improvement) compared to the benchmark while maintaining sensitivity constant at 85%.

**Figure 15:**
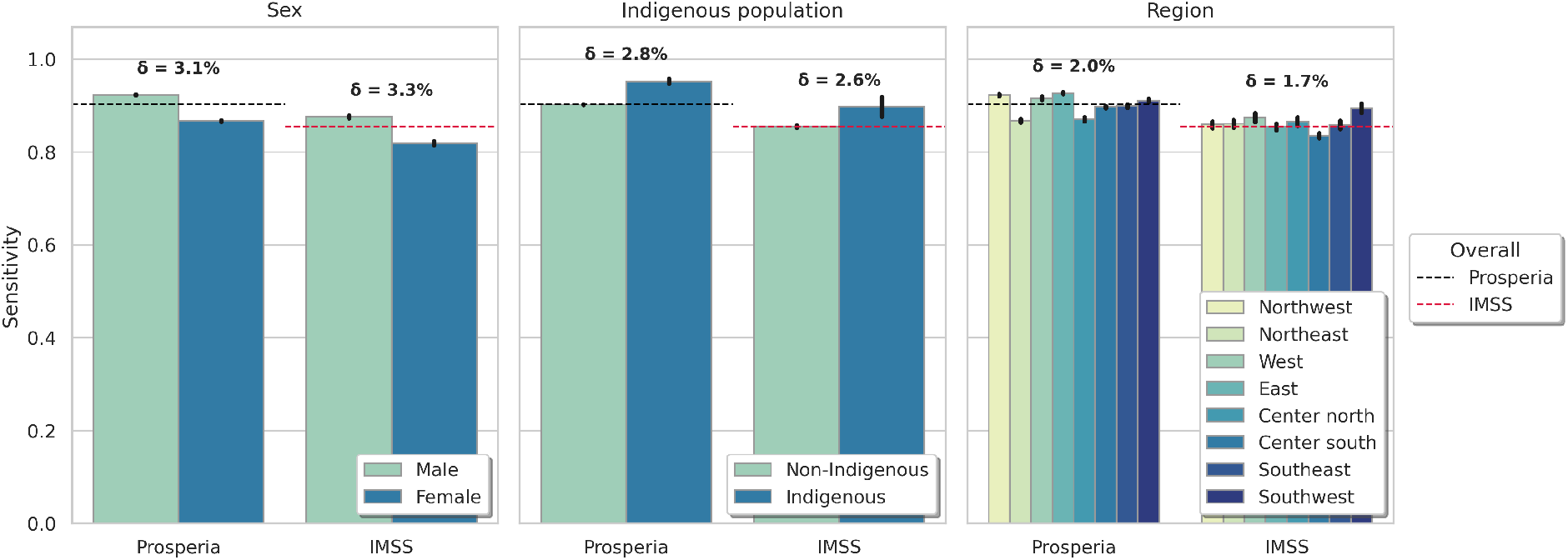
Equity across subgroups for the COVID-19 severity model and benchmark.

**Figure 16:**
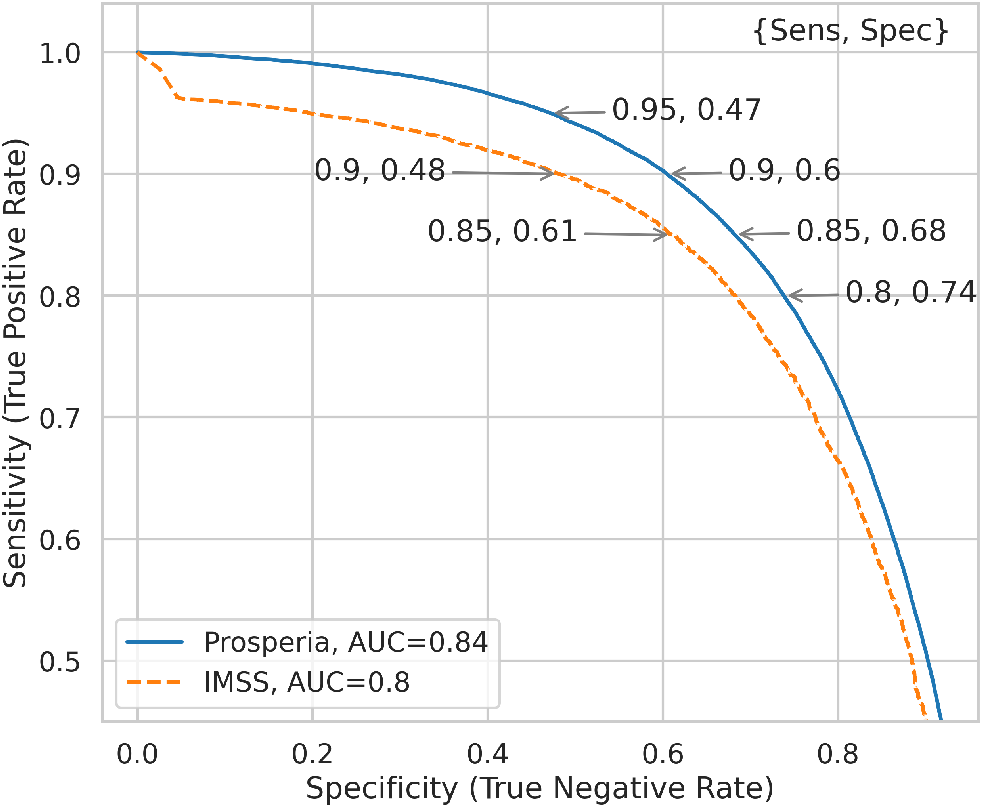
ROC curves of the COVID-19 severity model and benchmark.

#### Importance of features

The variable with the largest impact on the risk index was found to be the *age*. Other highly relevant variables include *state of residence, sex*, and comorbidities such as *diabetes, hypertension*, and *obesity* (Figure 17).

**Figure 17:**
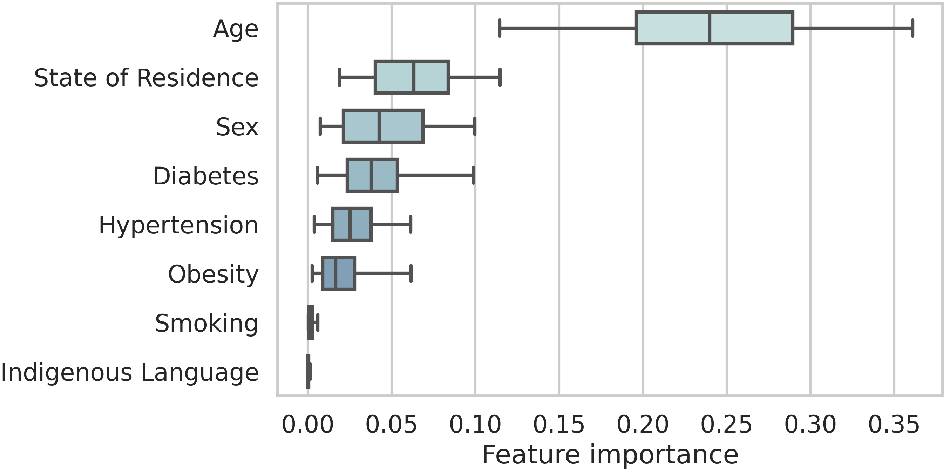
Importance of features for the COVID-19 severity model.

These high-relevance risk factors are in line with those found in most COVID-19 severity risk studies internationally [64, 13]. However, the *state of residence* variable was not studied or found in the aforecited studies. The relevance of this variable here found could be related to varying socioeconomic and quality of healthcare systems, as, for example, it is known that the fatality rate conditional on endotracheal intubation can vary widely across hospital networks [48], potentially due to differences in the quality of care. Finally, these results are in line with recommendations from Ghisolfi et al., on the importance of applying sub-national adjustments to COVID-19 IFR estimations [30].

#### Equity across subgroups

Both our model and the risk calculator developed by IMSS showed a very low disparity in all considered divisions, with the largest gap found across sex groups at only slightly over 3% (Figure 15).

## 5. Integrated Risk Assessment Platform

For public access at a large-scale, the risk models were deployed on a web and mobile platform and made accessible without cost through several institutional channels and social networks. The platform contains a short questionnaire, after which it provides a report with four risk indices: diabetes, hypertension, retinal damage due to diabetes or hypertension, and risk of severe COVID-19 complications. The questionnaire adapts to the answers of the user to maximize the certainty of the results. If the user has a medical diagnosis of diabetes or hypertension, the questionnaire expands to include questions regarding the progression and treatment of each disease.

Lastly, the report includes a report containing the risk indices, as well as information and references about health education on chronic diseases and their prevention, and contextualized links to public and private health institutions. The report is sent by email or text message to the users. Figures 18 and 19 show the visual appearance of the integrated platform’s questionnaire and report.

**Figure 18:**
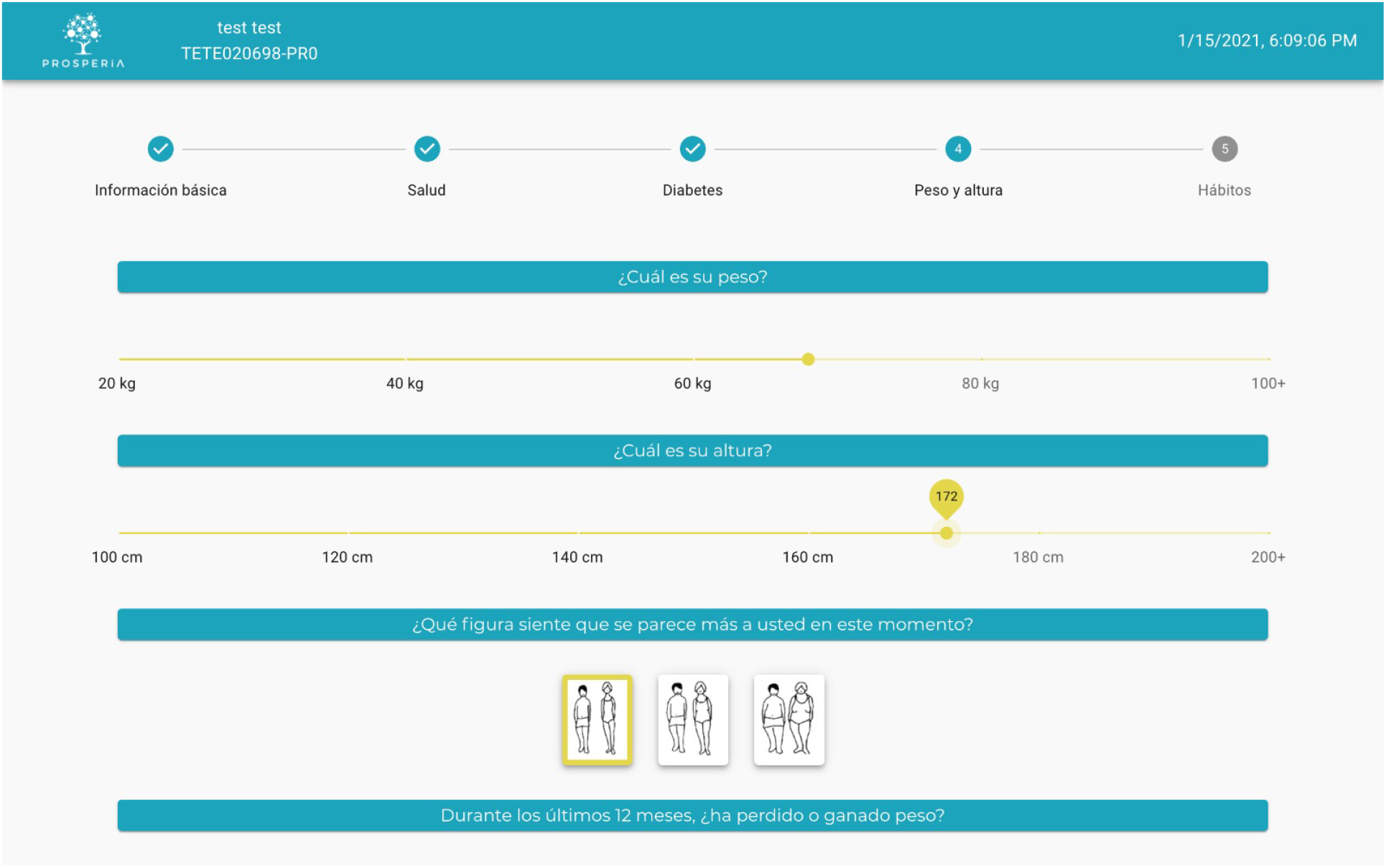
Questionnaire

**Figure 19:**
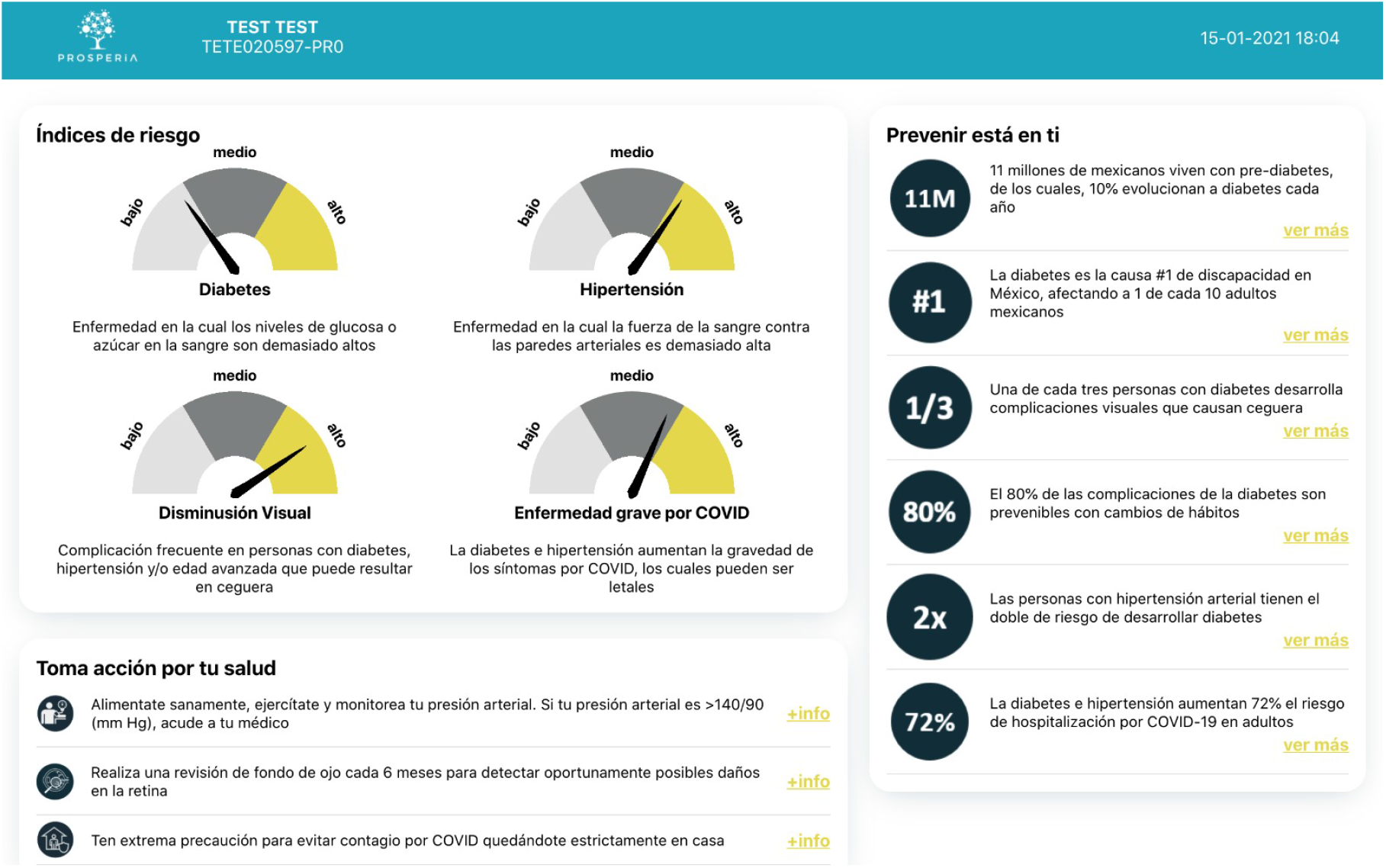
Results report

## 6. Conclusions

The use of large databases representative of the Mexican population, coupled with modern statistical learning methods, allowed the development of risk models with state-of-the-art accuracy and equity for two of the most relevant chronic diseases, their eye complications, and COVID-19 severity. Hence, these results highlight a) the relevance of collecting local, high-quality, public health surveys like ENSANUT, to assess and develop contextualized risk assessment tools; and b) the potential accuracy edge provided by statistical learning algorithms.

Similarly, this work highlights the relevance of assessing the accuracy of risk models in terms of algorithmic equity, beyond general population averages. Here we found that substantial disparities exist in several benchmark models currently in use, for subgroups defined by sex, indigenous origin, and geographical regions. In this vein, a contribution of the present work consists of showing that more accurate models based on the aforementioned characteristics can improve overall accuracy as well as equity across relevant subgroups.

Finally, the tools here presented can have a meaningful impact towards democratizing early detection of chronic diseases and their complications, by fostering public awareness and enabling large-scale preventive strategies in low-resource health systems, with the potential to ultimately raise social well-being.

## Data Availability

All data used in the present work is publicly available, and the corresponding references to data sources are included in the manuscript

## Acknowledgements

We are deeply thankful for the support of the EmpatIA initiative^8^ and its member organizations: the Inter-American Development Bank (IDB), the International Development Research Centre (IDRC), the Centro Latam Digital, and the Latin American Initiative for Open Data (ILDA). We are also thankful to Andrew Beckham for his valuable research assistantship.

## A. Appendix

## Footnotes

1 See this large repository of risk calculators for example: https://qxmd.com/calculate.

2 We used a linguistic criterion to determine indigenous origin as is usually done in Mexico [20]

3 Calculator available at: https://www.diabetes.org/risk-test

4 Prior to comparison, we applied a piece-wise linear function to the output of each model, to balance their sensitivity and specificity at two discrimination thresholds corresponding to medium and high risk.

5 We used a linguistic criterion to determine indigenous origin as is usually done in Mexico [20]

6 We used a linguistic criterion to determine indigenous origin as is usually done in Mexico [20]

7 Calculator available at: http://www.imss.gob.mx/covid-19/calculadora-complicaciones

8 EmpatIA: https://empatia.la/

## References

[1] Country profiles: Mexico. institute for health metrics and evaluation. http://www.healthdata.org/ mexico. Accessed: 2020-11-30.

[2] Instituto mexicano para la competitividad (imco). kilos de más, pesos de menos. el costo de la obesidad en méxico. 2015. https://imco.org.mx/wp-content/uploads/2015/01/20150127_ObesidadEnMexico_DocumentoCompleto.pdf. Accessed: 2020-12-10.

[3] Secretaría de salud. guía de práctica clínica, diagnóstico y tratamiento de retinopatía diabética. 2015. http://www.cenetec.salud.gob.mx/descargas/gpc/CatalogoMaestro/171_GPC_RETINOPATIA_DIABETICA/Imss_171RR.pdf. Accessed: 2020-12-10.

[4] Summary of revisions: Standards of medical care in diabetes—2020. Diabetes Care, 43(Supplement 1):S4–S6, December 2019.

[5] A. Abbasi, L. Peelen, E. Corpeleijn, Y. T. van der Schouw, R. Stolk, A. Spijkerman, A D. vander, K. Moons, G. Navis, S. Bakker, and J. Beulens. Prediction models for risk of developing type 2 diabetes: systematic literature search and independent external validation study. The BMJ, 345, 2012.

[6] André Altmann, Laura Toloşi, Oliver Sander, and Thomas Lengauer. Permutation importance: a corrected feature importance measure. Bioinformatics, 26(10):1340–1347, 04 2010.

[7] P. Aschner, Ramfis Nieto-Martínez, Alejandro Marin, and M. Ríos. Validation of the findrisk score as a screening tool for people with impaired glucose regulation in latin america using modified score points for waist circumference according to the validated regional cutoff values for abdominal obesity. Minerva Endocrinol, 37, 01 2012.

[8] Thor Aspelund, O Thornórisdóttir, Elin Olafsdottir, Arna Gudmundsdottir, Anna Einarsdottir, Jesper Mehlsen, S Einarsson, O Pálsson, G Einarsson, T Bek, and Einar Stefánsson. Individual risk assessment and information technology to optimise screening frequency for diabetic retinopathy. Diabetologia, 54:2525–32, 07 2011.

[9] Heejung Bang. Development and validation of a patient self-assessment score for diabetes risk. Annals of Internal Medicine, 151(11):775, December 2009.

[10] A. Barceló, A. Arredondo, A. Gordillo-Tobar, J. Segovia, and Anthony Qiang. The cost of diabetes in latin america and the caribbean in 2015: Evidence for decision and policy makers. Journal of Global Health, 7, 2017.

[11] A. Bernabé-Ortiz, P. Perel, J. Miranda, and L. Smeeth. Diagnostic accuracy of the finnish diabetes risk score (findrisc) for undiagnosed t2dm in peruvian population. Primary Care Diabetes, 12:517 – 525, 2018.

[12] M Bhargava, M K Ikram, and T Y Wong. How does hypertension affect your eyes? Journal of Human Hypertension, 26(2):71–83, April 2011.

[13] Stephanie Bialek, Ellen Boundy, Virginia Bowen, Nancy Chow, Amanda Cohn, Nicole Dowling, Sascha Ellington, Ryan Gierke, Aron Hall, Jessica MacNeil, Priti Patel, Georgina Peacock, Tamara Pilishvili, Hilda Razzaghi, Nia Reed, Matthew Ritchey, and Erin Sauber-Schatz and. evere outcomes among patients with coronavirus disease 2019 (COVID-19) — united states, february 12–march 16, 2020. MMWR. Morbidity and Mortality Weekly Report, 69(12):343–346, March 2020.

[14] Carissa Bonner, M. A. Fajardo, Samuel Hui, Renee Stubbs, and L. Trevena. Clinical validity, understandability, and actionability of online cardiovascular disease risk calculators: Systematic review. Journal of Medical Internet Research, 20, 2018.

[15] S. Borzouei and A. Soltanian. Application of an artificial neural network model for diagnosing type 2 diabetes mellitus and determining the relative importance of risk factors. Epidemiology and Health, 40, 2018.

[16] Leo Breiman. Random forests. Machine Learning, 45(1):5–32, 2001.

[17] R. Carrillo-Larco, D. J. Aparcana-Granda, J. R. Mejía, N. Barengo, and A. Bernabé-Ortiz. Risk scores for type 2 diabetes mellitus in latin america: a systematic review of population-based studies. Diabetic Medicine, 36:1573 – 1584, 2019.

[18] D Cavan, L Makaroff, J da Rocha Fernandes, M Sylvanowicz, P Ackland, J Conlon, D Chaney, A Malhi, and J Barratt. The diabetic retinopathy barometer study: Global perspectives on access to and experiences of diabetic retinopathy screening and treatment. Diabetes research and clinical practice, 129:16—24, July 2017.

[19] Consejo Nacional de Ciencia y Tecnología. Covid-19 tablero méxico. https://datos.covid-19.conacyt.mx/, 2020. Accessed: 2020-11-24.

[20] Instituto Nacional de los Pueblos Indígenas. Indicadores Socioeconómicos de los Pueblos Indígenas de México, 2015.

[21] Instituto Mexicano del Seguro Social. Diseña IMSS calculadora para evaluar nivel de gravedad a la salud en caso de padecer COVID-19.

[22] Edgar Denova-Gutiérrez, Hugo Lopez-Gatell, Jose L. Alomia-Zegarra, Ruy López-Ridaura, Christian A. Zaragoza-Jimenez, Dwigth D. Dyer-Leal, RicardoCortés-Alcala, Tania Villa-Reyes, Rosaura Gutiérrez-Vargas, Kathia Rodríguez-González, Carlos Escondrillas-Maya, Tonatiuh Barrientos-Gutiérrez, Juan A. Rivera, and Simón Barquera. The association of obesity, type 2 diabetes, and hypertension with severe coronavirus disease 2019 on admission among mexican patients. Obesity, 28(10), October 2020.

[23] Diabetes Prevention Program Research Group, David M. Nathan, Elizabeth Barrett-Connor, Jill P. Crandall, Sharon L. Edelstein, Ronald B. Goldberg, Edward S. Horton, William C. Knowler, Kieren J. Mather, Trevor J. Orchard, Xavier Pi-Sunyer, David Schade, and Marinella Temprosa. Long-term effects of lifestyle intervention or metformin on diabetes development and microvascular complications over 15-year follow-up: The diabetes prevention program outcomes study. The Lancet Diabetes and Endocrinology, 3(11):866–875, November 2015. Publisher Copyright: © 2015 Elsevier Ltd. Copyright: Copyright 2017 Elsevier B.V., All rights reserved.

[24] Dirección General de Epidemiología. Covid-19. Technical report, Secretaría de Salud, 2020.

[25] Bradley Efron and Robert Tibshirani. An introduction to the bootstrap. Number 57 in Monographs on statistics and applied probability. Chapman & Hall, New York, 1993.

[26] Antonio Eleuteri, Anthony C. Fisher, Deborah M. Broadbent, Marta García-Fiñana, Christopher P. Cheyne, Amu Wang, Irene M. Stratton, Mark Gabbay, Daniel Seddon, and Simon P. Harding. Individualised variable-interval risk-based screening for sight-threatening diabetic retinopathy: the liverpool risk calculation engine. Diabetologia, 60(11):2174–2182, August 2017.

[27] Tom Fawcett. An introduction to ROC analysis. Pattern Recognition Letters, 27(8):861–874, June 2006.

[28] International Diabetes Federation. Idf diabetes atlas, 9th edn. https://www.diabetesatlas.org, 2019. Accessed: 2021-01-04.

[29] Jerome H. Friedman. Greedy function approximation: A gradient boosting machine. The Annals of Statistics, 29(5):1189–1232, October 2001.

[30] Selene Ghisolfi, Ingvild Almås, Justin C Sandefur, Tillman von Carnap, Jesse Heitner, and Tessa Bold. Predicted COVID-19 fatality rates based on age, sex, comorbidities and health system capacity. BMJ Global Health, 5(9), 2020. Publisher: BMJ Specialist Journals _eprint: https://gh.bmj.com/content/5/9/e003094.full.pdf.

[31] C. Glumer, B. Carstensen, A. Sandbaek, T. Lauritzen, T. Jorgensen, and K. Borch-Johnsen. A danish diabetes risk score for targeted screening: The inter99 study. Diabetes Care, 27(3):727–733, February 2004.

[32] Paul Grant. Management of diabetes in resource-poor settings. Clinical Medicine, 13(1):27–31, February 2013.

[33] Trevor Hastie, Jonathan Taylor, Robert Tibshirani, and Guenther Walther. Forward stagewise regression and the monotone lasso. Electronic Journal of Statistics, 1(0):1–29, 2007.

[34] Trevor Hastie, Robert Tibshirani, and Jerome Friedman. The Elements of Statistical Learning. Springer Series in Statistics. Springer New York, New York, NY, 2009.

[35] Kenneth E. Heikes, David M. Eddy, Bhakti Arondekar, and Leonard Schlessinger. Diabetes risk calculator. Diabetes Care, 31(5):1040–1045, 2008.

[36] W. Herman, P. Smith, T. J. Thompson, M. Engelgau, and R. Aubert. A new and simple questionnaire to identify people at increased risk for undiagnosed diabetes. Diabetes Care, 18:382 – 387, 1995.

[37] William H. Herman, Sharon L. Edelstein, Robert E. Ratner, Maria G. Montez, Ronald T. Ackermann, Trevor J. Orchard, Mary A. Foulkes, Ping Zhang, Christopher D. Saudek, and Morton B. Brown. The 10-year cost-effectiveness of lifestyle intervention or metformin for diabetes prevention: An intent-to-treat analysis of the dpp/dppos. Diabetes Care, 35(4):723–730, April 2012. Copyright: Copyright 2012 Elsevier B.V., All rights reserved.

[38] I. Kakadiaris, Michalis Vrigkas, A. Yen, T. Kuznetsova, M. Budoff, and M. Naghavi. Machine learning outperforms acc/aha cvd risk calculator in mesa. Journal of the American Heart Association: Cardiovascular and Cerebrovascular Disease, 7, 2018.

[39] Abhijit V. Kshirsagar, Ya lin Chiu, Andrew S. Bom-back, Phyllis A. August, Anthony J. Viera, Romulo E. Colindres, and Heejung Bang. A hypertension risk score for middle-aged and older adults. The Journal of Clinical Hypertension, 12(10):800– 808, July 2010.

[40] Hugo Larochelle, Dumitru Erhan, Aaron Courville, James Bergstra, and Yoshua Bengio. An empirical evaluation of deep architectures on problems with many factors of variation. In Proceedings of the 24th international conference on Machine learning - ICML ‘07. ACM Press, 2007.

[41] Wenhua Liang, Hengrui Liang, Limin Ou, Binfeng Chen, Ailan Chen, Caichen Li, Yimin Li, Weijie Guan, Ling Sang, Jiatao Lu, Yuanda Xu, Guo-qiang Chen, Haiyan Guo, Jun Guo, Zisheng Chen, Yi Zhao, Shiyue Li, Nuofu Zhang, Nanshan Zhong, and Jianxing He and. Development and validation of a clinical risk score to predict the occurrence of critical illness in hospitalized patients with COVID-19. JAMA Internal Medicine, 180(8):1081, August 2020.

[42] Jaana Lindström and Jaakko Tuomilehto. The diabetes risk score: a practical tool to predict type 2 diabetes risk. Diabetes care, 26(3):725—731, March 2003.

[43] D.G. Lugo-Palacios and J. Cairns. The financial and health burden of diabetic ambulatory care sensitive hospitalisations in mexico. Salud Publica Mex, 58(1):33–40, Jan-Feb 2016.

[44] F. Márquez-Celedonio, Obdulia Téxon-Fernández, A. Chávez-Negrete, Sergio Hernández-López, Sadoc Marín-Rendón, and Susana Berlín-Lascurain. [clinical effect of lifestyle modification on cardiovascular risk in prehypertensives: Prehiper i study]. Revista espanola de cardiologia, 62 1:86–90, 2009.

[45] Mathematica. 19 and me: A covid-19 risk calculator. https://19andme.covid19.mathematica. org/, 2020. Accessed: 2020-09-04.

[46] Kathryn L. McCance and Sue E. Huether, editors. Pathophysiology: the biologic basis for disease in adults and children. Elsevier, St. Louis, Missouri, eighth edition edition, 2019.

[47] Kenny Mendoza-Herrera, Amado D. Quezada, Andrea Pedroza-Tobías, Cesar Hernández-Alcaraz, Jans Fromow-Guerra, and Simón Barquera. A diabetic retinopathy screening tool for low-income adults in mexico. Preventing Chronic Disease, 14, October 2017.

[48] Silvio A. Ñamendys-Silva, AlanGutiérrez-Villaseñor, and Juan P. Romero-González. Hospital mortality in mechanically ventilated COVID-19 patients in mexico. Intensive Care Medicine, 46(11):2086–2088, September 2020.

[49] Ramfis Nieto-Martínez, Juan P. González-Rivas, Pablo Aschner, Noël C. Barengo, and Jeffrey I. Mechanick. Transculturalizing diabetes prevention in latin america. Annals of Global Health, 83(3-4):432, August 2017.

[50] D. Noble, R. Mathur, T. Dent, C. Meads, and T. Greenhalgh. Risk models and scores for type 2 diabetes: systematic review. The BMJ, 343, 2011.

[51] Omolola I. Ogunyemi, Meghal Gandhi, and Chandler Tayek. Predictive Models for Diabetic Retinopathy from Non-Image Teleretinal Screening Data. AMIA Joint Summits on Translational Science proceedings. AMIA Joint Summits on Translational Science, 2019:472–477, 2019.

[52] Elvis Ojaimi, Thanh T. Nguyen, Ronald Klein, F.M. Amirul Islam, Mary Frances Cotch, Barbara E.K. Klein, Jie-Jin Wang, and Tien Yin Wong. Retinopathy signs in people without diabetes. Ophthalmology, 118(4):656–662, April 2011.

[53] Markus Ojala and Gemma C. Garriga. Permutation tests for studying classifier performance. In 2009 Ninth IEEE International Conference on Data Mining. IEEE, December 2009.

[54] World Health Organization. Hypertension. https://www.who.int/news-room/fact-sheets/detail/hypertension, 2010. Accessed: 2021-01-04.

[55] World Health Organization. Coronavirus. https://www.who.int/health-topics/coronavirus, 2020. Accessed: 2020-09-20.

[56] World Health Organization. Who coronavirus disease (covid-19) dashboard. https://covid19.who.int/, 2020. Accessed: 2021-01-04.

[57] Sajida Perveen, M. Shahbaz, K. Keshavjee, and A. Guergachi. Prognostic modeling and prevention of diabetes using machine learning technique. Scientific Reports, 9, 2019.

[58] E. Puentes-Rosas, S. Sesma, and O. Gómez-Dantés. Estimación de la población con seguro de salud en méxico mediante una encuesta nacional. Salud Pública de México, 2015.

[59] Germán Rodríguez. Lecture notes on generalized linear models. https://data.princeton.edu/ wws509/notes/, 2007.

[60] T. Shamah-Levy, E. Vielma-Orozco, O. Heredia-Hernández, M. Romero-Martínez, J. Mojica-Cuevas, L. Cuevas-Nasu, J. A. Santaella-Castell, and J. Rivera-Dommarco. National survey of health and nutrition 2018-19. national results [encuesta nacional de salud y nutrición 2018-19. resultados nacionales]. Technical report, Instituto Nacional de Salud Pública, 2020.

[61] Robert Tibshirani. Regression shrinkage and selection via the lasso. Journal of the Royal Statistical Society: Series B (Methodological), 58(1):267–288, January 1996.

[62] Hsin-Yi Tsao, Pei-Ying Chan, and Emily Chia-Yu Su. Predicting diabetic retinopathy and identifying interpretable biomedical features using machine learning algorithms. BMC Bioinformatics, 19(S9):283, August 2018.

[63] Patrick G. T. Walker, Charles Whittaker, Oliver J Watson, Marc Baguelin, Peter Winskill, Arran Hamlet, Bimandra A. Djafaara, Zulma Cucunubá, Daniela Olivera Mesa, Will Green, Hayley Thompson, Shevanthi Nayagam, Kylie E. C. Ainslie, Sangeeta Bhatia, Samir Bhatt, Adhiratha Boonyasiri, Olivia Boyd, Nicholas F. Brazeau, Lorenzo Cattarino, Gina Cuomo-Dannenburg, Amy Dighe, Christl A. Donnelly, Ilaria Dorigatti, Sabine L. van Elsland, Rich FitzJohn, Han Fu, Katy A.M. Gaythorpe, Lily Geidelberg, Nicholas Grassly, David Haw, Sarah Hayes, Wes Hinsley, Natsuko Imai, David Jorgensen, Edward Knock, Daniel Laydon, Swapnil Mishra, Gemma Nedjati-Gilani, Lucy C. Okell, H. Juliette Unwin, Robert Verity, Michaela Vollmer, Caroline E. Walters, Haowei Wang, Yuanrong Wang, Xiaoyue Xi, David G Lalloo, Neil M. Ferguson, and Azra C. Ghani. The impact of COVID-19 and strategies for mitigation and suppression in low- and middle-income countries. Science, page eabc0035, June 2020.

[64] Elizabeth J. Williamson, Alex J. Walker, Krishnan Bhaskaran, Seb Bacon, Chris Bates, Caroline E. Morton, Helen J. Curtis, Amir Mehrkar, David Evans, Peter Inglesby, Jonathan Cockburn, Helen I. McDonald, Brian MacKenna, Laurie Tomlinson, Ian J. Douglas, Christopher T. Rentsch, Rohini Mathur, Angel Y. S. Wong, Richard Grieve, David Harrison, Harriet Forbes, Anna Schultze, Richard Croker, John Parry, Frank Hester, Sam Harper, Rafael Perera, Stephen J. W. Evans, Liam Smeeth, and Ben Goldacre. Factors associated with COVID-19-related death using OpenSAFELY. Nature, 584(7821):430–436, July 2020.

[65] Tien Y. Wong and Paul Mitchell. Hypertensive retinopathy. New England Journal of Medicine, 351(22):2310–2317, November 2004.

[66] J. W. Y. Yau, S. L. Rogers, R. Kawasaki, E. L. Lamoureux, J. W. Kowalski, T. Bek, S.-J. Chen, J. M. Dekker, A. Fletcher, J. Grauslund, S. Haffner, R. F. Hamman, M. K. Ikram, T. Kayama, B. E. K. Klein, R. Klein, S. Krishnaiah, K. Mayurasakorn, J. P. O’Hare, T. J. Orchard, M. Porta, M. Rema, M. S. Roy, T. Sharma, J. Shaw, H. Taylor, J. M. Tielsch, R. Varma, J. J. Wang, N. Wang, S. West, L. Xu, M. Yasuda, X. Zhang, P. Mitchell, and T. Y. Wong and. Global prevalence and major risk factors of diabetic retinopathy. Diabetes Care, 35(3):556–564, February 2012.

